# Patterns of 2,4-Dinitrophenol Use as Discussed on Social Media

**DOI:** 10.1101/2020.04.26.20080382

**Authors:** Michael Chary, Karun Ellango, Michele Burns

**Author notes:** Corresponding author (Michael Chary).

## Abstract

**Importance:** The usage of social media is associated with worsening perceptions of body image and increasing access to, and use of, toxic weight loss supplements. Little is known about the effect of nonlethal doses of one mechanistically unique supplement, 2,4-dinitrophenol (DNP). DNP has been banned by the FDA making human studies difficult, but the public still consumes DNP leading to a gap in our knowledge on the effects of DNP. Here we use social media to investigate the use of DNP, providing the largest characterization of its usage to date.

**Objective:** Determine the doses of DNP generally consumed, adverse effects at those doses, and coingestants.

**Design:** Cross-sectional analysis of Internet posts.

**Setting:** Our study collected publicly available data from 2017–2018 from Internet discussion forums (also called bulletin boards) dedicated to the discussion of weight loss and body building.

**Participants:** Participants are anonymous posters on these Internet forums.

**Main Measures:** Our main measure was the distribution of reported doses of DNP consumed. Our secondary measure was the frequency of adverse effects reported at those doses.

**Results:** We collected 661 posts across 5 online forums. The most commonly ingested dose reported was 150 mg (1-2 pills, depending on formulation), followed by 300 mg (2-3 pills). The most commonly reported adverse effects were sweating and a sensation of warmth, followed by yellow discoloration of secretions. The most common coingestants were antihistamines, cetirizine and loratadine.

**Conclusions and Relevance:** 2,4-dinitrophenol is a mechanistically unique weight loss agent reported to be associated with sweating and a sensation of warmth at the most commonly reported ingested doses. Common co-ingestants are antihistamines, although itching was not directly reported as a side effect. Coingestion of an antihistamine, which can lessen the body’s ability to dissipate heat, could worsen the side effects of DNP. This is the first formal description derived from social media of DNP usage at nonlethal doses. Further investigation is needed to determine the therapeutic index of DNP. Less toxic derivatives may provide a starting point for pharmacological adjuncts to weight-loss.

## INTRODUCTION

The use of social media is associated with worsening perceptions of body image^1^, and may increase interest in substance use to lose fat among those suffering from eating disorders or body dysmorphic disorders as well as fitness enthusiasts. Those seeking to increase lean muscle mass may alternate periods of *bulking*, heavy training with a caloric surplus to gain overall body mass, with periods of *cutting*, training with a caloric deficit to reduce adipose tissue^2^. The ideal bulking phase builds only lean body mass (muscle). The ideal cutting phase decreases only adipose tissue and preserves lean body mass. Anabolic steroids^3^, prescription medications including weight loss agents^4^, and nutritional supplements^5^ have been used to augment the physiologic response to each phase.

The Food and Drug Administration (FDA) has approved 5 medications to aid weight loss, orlistat (lipase inhibitor), phentermine (phenethylamine)/topiramate (anticonvulsant), lorcaserin (central serotonin 2C agonist), naltrexone (opioid partial agonist)/bupropion (stimulant), and liraglutide (glucagon-like peptide-1 agonist). These medications can lead to 3-5 kg net weight loss at one year^6^. 2,4-dinitrophenol (DNP) can be an attractive alternative to those trying to rapidly lose weight as DNP, anecdotally, can lead to 3-5 kg of weight loss *each week^7^*. Use of DNP is associated with adverse effects, ranging from cataracts to death from hyperthermia and cardiovascular collapse^8^.

DNP uncouples oxidative phosphorylation by dissipating the proton gradient across the inner mitochondrial membrane^9^. It was first prescribed by physicians for weight loss in 1933^10,11^. DNP was removed from the market in 1938 and labeled by the FDA as not fit for human consumption after reports of fatal hyperthermia and progressive cataracts^12^. It is currently approved in the United States only as an industrial pesticide.

Despite the ban by the FDA and its toxicity, DNP use seems to be increasing. Of the 25 reports of fatalities, 12 occurred after 2000^7^. Of those 12 reports, 7 involved young athletic men taking DNP to reduce body fat without losing muscle mass.

There is a gap in our knowledge of the effects and toxicity of DNP at doses that do not lead to cardiovascular collapse and medical intervention. It is unethical to administer a substance the FDA has banned for human consumption. The public, despite this ban, consumes DNP in search of more effective pharmaceutical adjuncts for weight loss and records their experiences on social media. Our prior work demonstrated that natural language processing of narrative text from social media can identify dose-effect relationships^13^.

The purpose of this study is to investigate the usage of 2,4-dinitrophenol (DNP) as reported on social media in order to better understand the unintended effects of DNP at nonlethal doses. We analyzed online discussion forums to estimate dosage, effects, and patterns of co-ingestion.

## METHODS

All software was written in Python by authors MC and KE. All analysis was conducted by MC. Figure 1 outlines the study flow. Throughout this section lowercase italic text refers to a specific Python library, *e.g. scipy* is a Python library that provides scientific computation functions. We retrieved text from 5 online forums (Figure 1, upper left), used natural language processing to extract posts mentioning substances (Figure 1, middle left) and the subset of those posts that mentioned only DNP (Figure 1, lower left). From the posts mentioning at least one substance we calculated the correlation of appearances between substances (middle right). From the posts mentioning only DNP, we calculated how often each dose was mentioned and the effects mentioned at each dosage (lower right).

**Figure 1.**
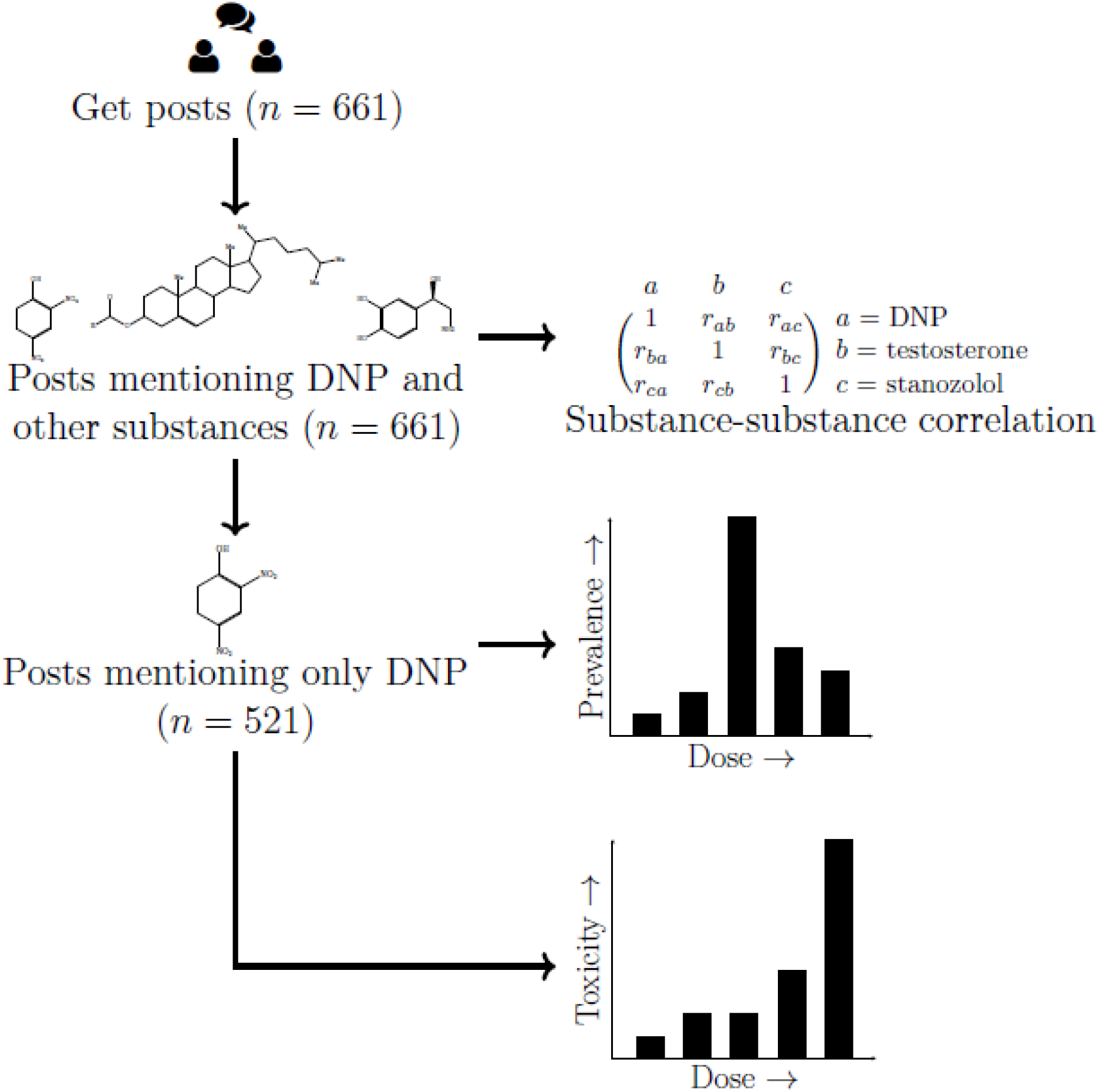
Schematic of study flow.

### Acquisition of Data

Publicly available posts that contained lexical or semantic variants of DNP (2,4-dinitrophenol, DNP, and all spelling variants) were retrieved from the websites listed in Table 1. These posts were retrieved and converted from HTML into plain text using the *scrapy* module in Python^14^.

### Identification of mentions of DNP and Co-ingestants

We used *nltk^15^ to* label each word in each post by its most likely part-of-speech (*e.g*. noun, adjective, *etc*.). We identified candidate substances as words that were tagged as nouns or abbreviations. Author MC manually reviewed these 252 candidate words to identify all unique names of substances construct a mapping from all lexical and semantic of those names to standard nomenclature. Equation (1) provides an example mapping that directs posts that mention *Tren* or *trenbalone* be grouped together under the heading *trenbolone*. Trenbolone is an anabolic steroid and a derivative of 19-nortestosterone. One colloquial term for trenbolone is *tren*.

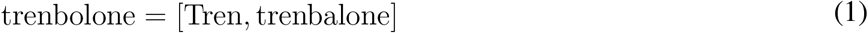

### Identification of doses

To extract doses we isolated posts that mentioned only DNP and one dose. We used *nltk* to identify and extract numbers from each post, whether written with numerals or written out. We then used *word2number* to convert numbers written in text into numerals, *e.g*. to convert *three* to 3. When dosing frequency was given, *e.g. 500 mg divided over twice a day*, author MC manually converted this to a single dose (*250 mg*).

### Correlation between mentions of substances

We created a mathematical representation of these data as a two-dimensional matrix, X. Each row in the matrix represented a post. Each column represented a substance. The *ij*th entry, where *i* refers to the row number and *j* refers to the column number, was *n* if the *i*th document mentioned the *j*th substance *n* times. For example, if the third post mentioned the second substance four times, then the entry at *X*_32_ = 4. We scaled each column in the matrix to have a norm of 1. We calculated the correlation between two words as the dot (inner) product of their corresponding vectors.

The ordering of posts and substances in the matrix is arbitrary and has no effect on subsequent analysis. See Chary et al. (2018)^16^ for more mathematical detail.

### Statistical significance of correlation

Bootstrapping provides a robust nonparametric way to estimate confidence intervals and *p*-values for statistical inference^17^. We used bootstrapping to calculate the statistical significance of substance-substance correlations. Briefly, we randomly shuffled the matrix described above 10,000 times and recalculated all correlation coefficients after each shuffle. To control for the comparison of multiple hypotheses we used the Benjamini-Hochberg correction^18^ with a false discovery rate set to 0.05.

## RESULTS

### Acquisition of Data

We retrieved 661 unique posts from 5 web forums (Table 1). The posts were posted online between Dec 2017 and Dec 2018. The most frequently mentioned substance is DNP as the forums were queried for posts that contained DNP or a DNP-related keyword. The number of posts mentioning DNP (661) is the same as the number of unique posts retrieved, providing an internal check that we retrieved only posts that mentioned DNP.

From these posts we identified 134 candidate names of co-mentioned substances. Figure 3 shows the 25 substances mentioned in the most posts. The remaining 24 substances relate to anabolism (testosterone, trenbolone, insulin, alpha alkylated steroid), weight loss and anabolism (clenbuterol), supplementation during DNP use (calcium, T3, sodium, Vitamin E, ethanol), or substances used to explain the effects of DNP by analogy (noradrenaline, epinephrine).

**Table 1.**
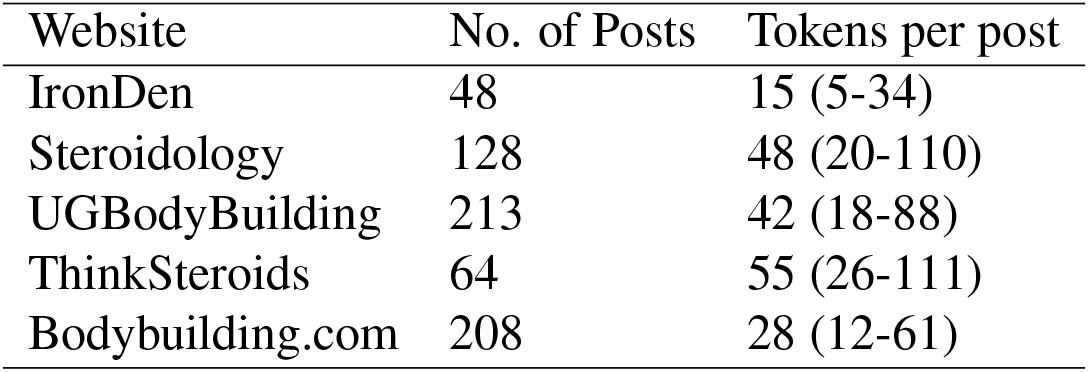
**Left column:** Name of web site, URL (Uniform Resource Locator) to main page of web site in parenthesis. **Middle column:** Number of posts from website that mention a DNP keyword. **Right column:** Median (interquartile range) of tokens (words, abbreviations, acronyms, see text for further explanation) in each post.

| Website | No. of Posts | Tokens per post |
| --- | --- | --- |
| IronDen | 48 | 15 (5-34) |
| Steroidology | 128 | 48 (20-110) |
| UGBodyBuilding | 213 | 42 (18-88) |
| ThinkSteroids | 64 | 55 (26-111) |
| Bodybuilding.com | 208 | 28 (12-61) |

**Figure 2.**
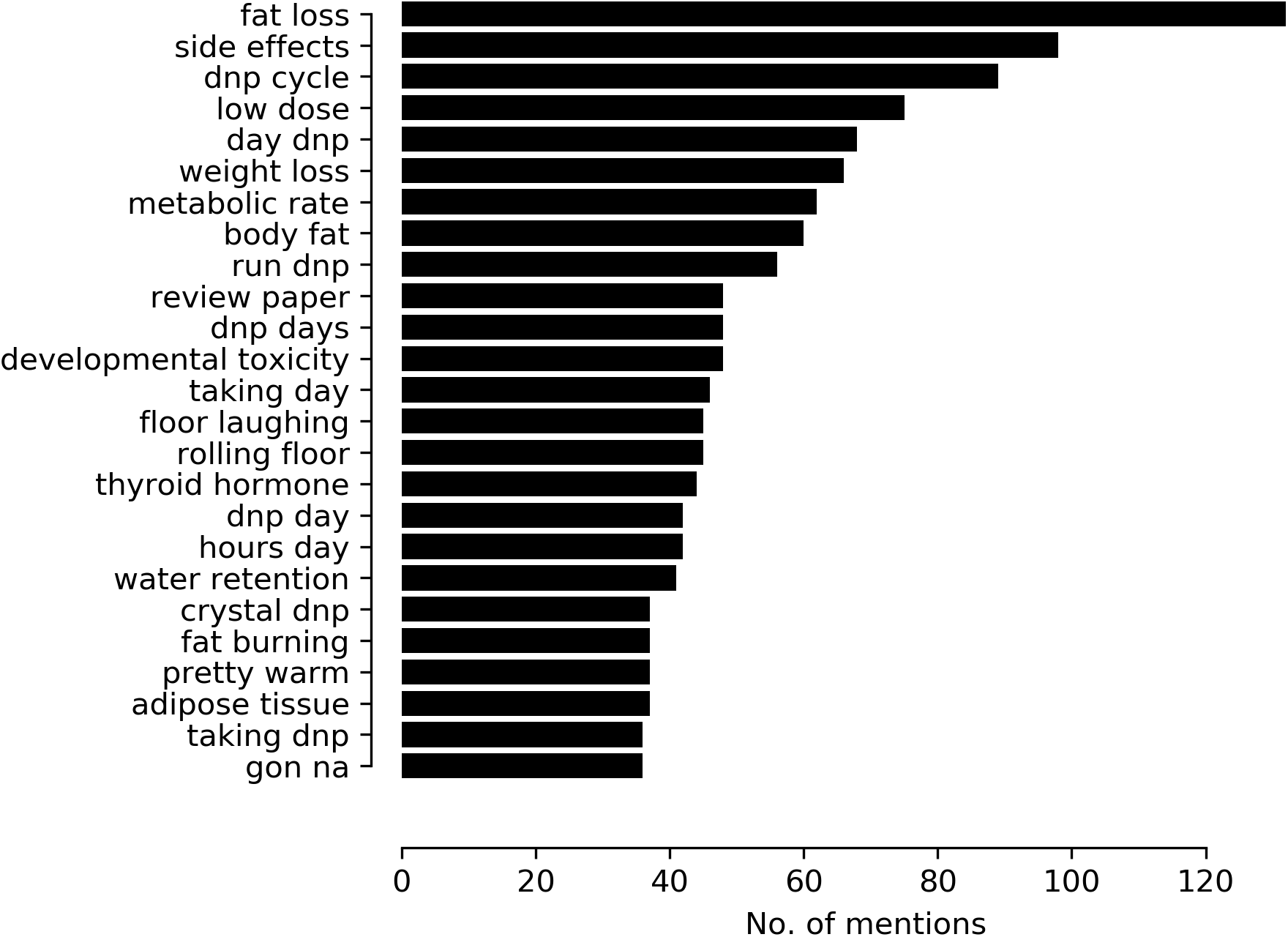
Twenty-five most frequently mentioned pairs of tokens (bigrams). Y-axis shows bigram. X-axis shows number of mentions across all posts mentioning DNP. Text shown after natural language processing but no copy-editing.

### Identification of doses

**Figure 3.**
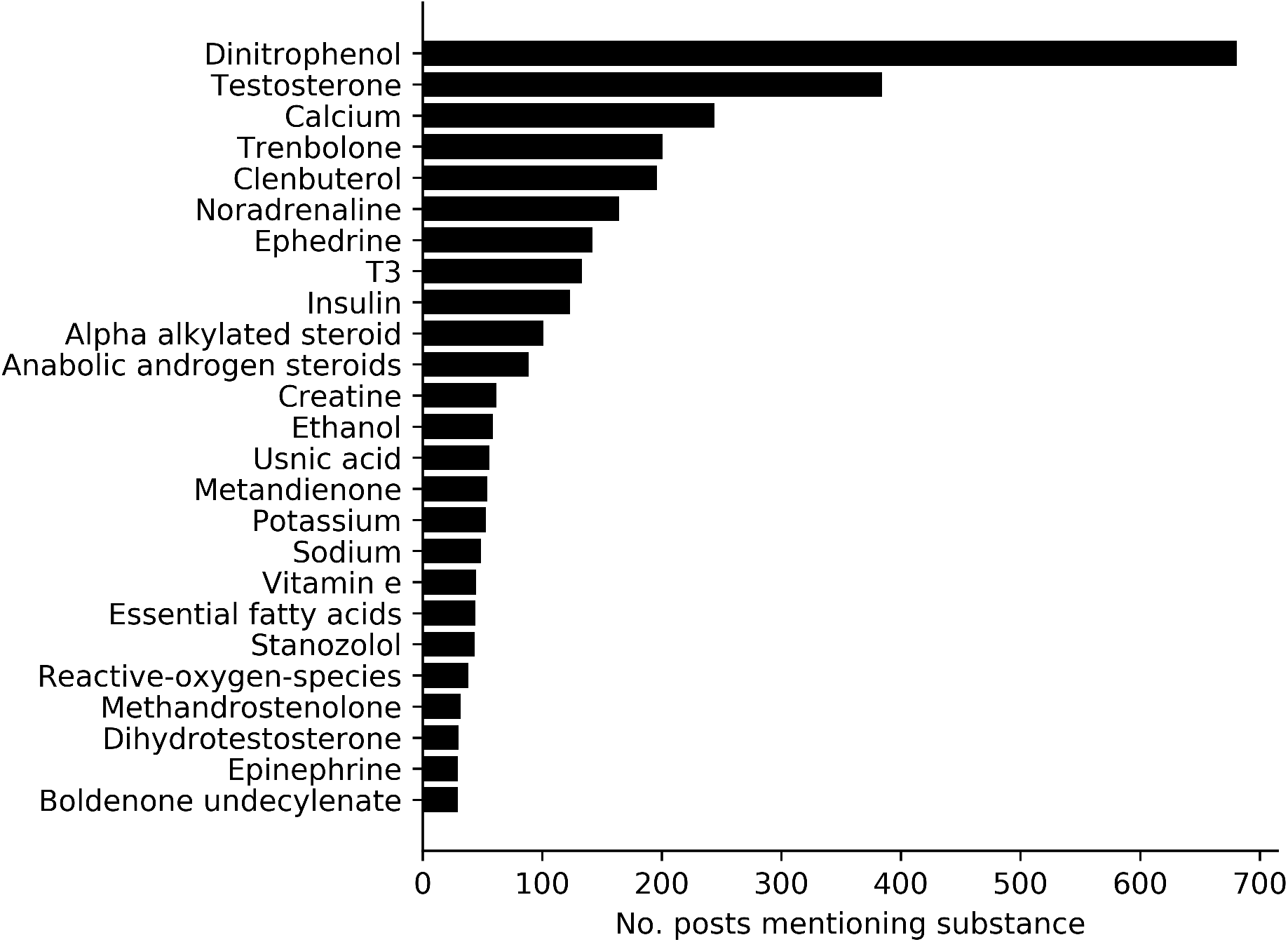
Distribution of substance mentions over all unique posts. Y-axis indicates name of substance; IUPAC or generic name used when applicable. X-axis indicates number of posts in which substance was mentioned.

To better understand the doses consumed by the general public we extracted the mentions of doses consumed from online discussions (Figure 4). The most common dosing was 150 mg twice a day, followed by 300 mg twice a day. The rightmost bar in Figure 4 refers to one reported ingestion of 3000 mg that was fatal.

DNP comes in two forms, crystalline or powder. The term *crystalline* denotes anhydrous DNP, which is expected to have more DNP per unit weight than the powder form. Crystalline DNP is sold in 100 mg pills. Powder DNP is sold in 150 mg pills. Both are also sold as lose “powders” that consumers can themselves pack into pills. Online posts did not consistently distinguish between crystalline and powder DNP. We chose bins of 150 mg to reflect the amount in one powder DNP pill. The most commonly reported doses correspond to 1–2 pills twice a day.

**Figure 4.**
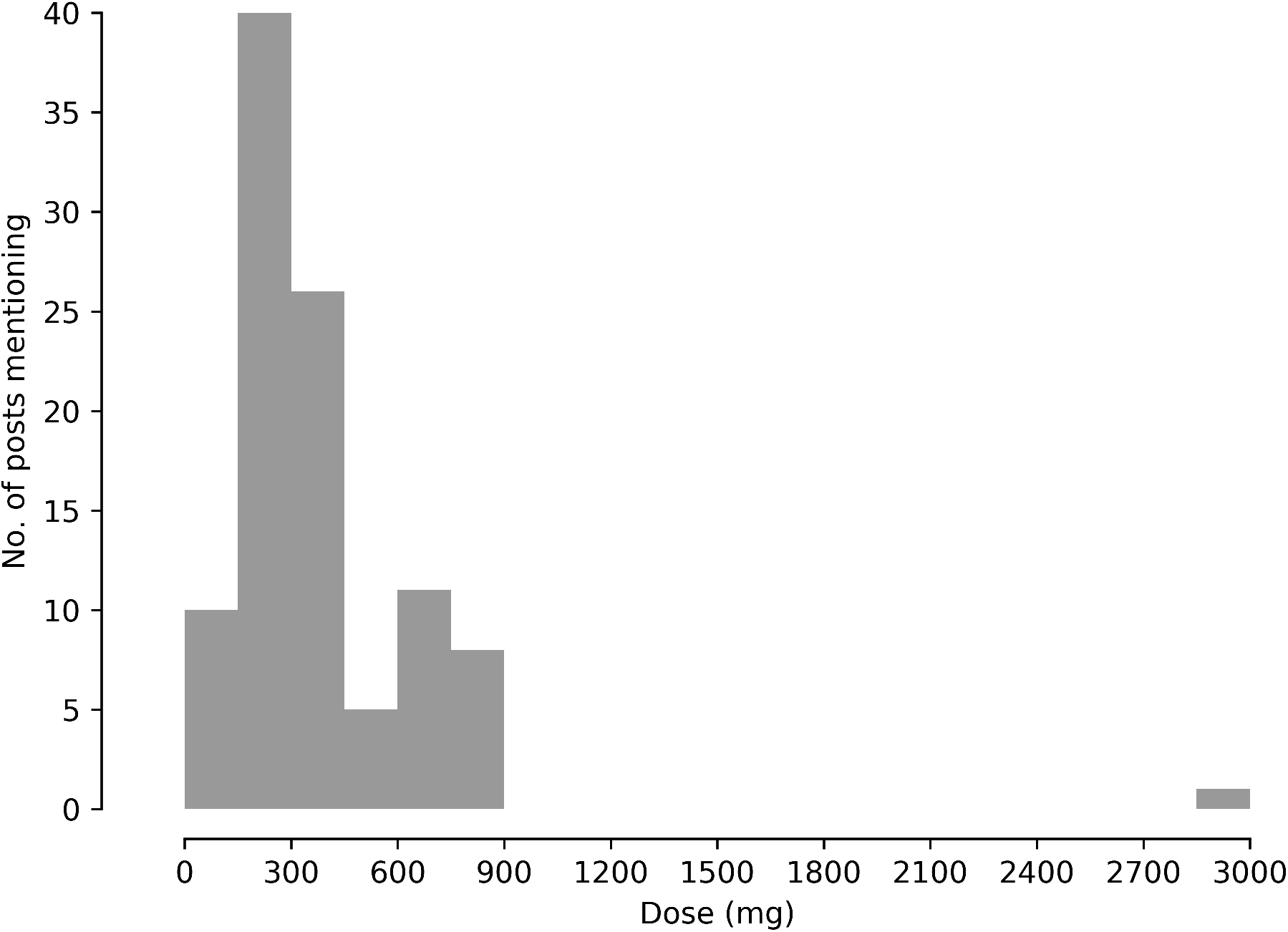
Distribution of individual doses in single DNP ingestion. X-axis shows mg per dose. Y-axis shows number of posts mentioning each dose. Bin size =150 mg.

### Identification of Effects

The most common words mentioned, after DNP, referred to the effects (fat, energy, feel, low, good) and timing (day, days, cycle, weeks) of taking DNP (Figure 5). To better understand the linguistic content, we calculated all pairs of tokens (Figure 2). The pairs of words that occurred together statistically significantly more than chance also described the effects (fat loss, weight loss, fat burning, pretty warm) and timing (dnp cycle, dnp days, low dose).

**Figure 5.**
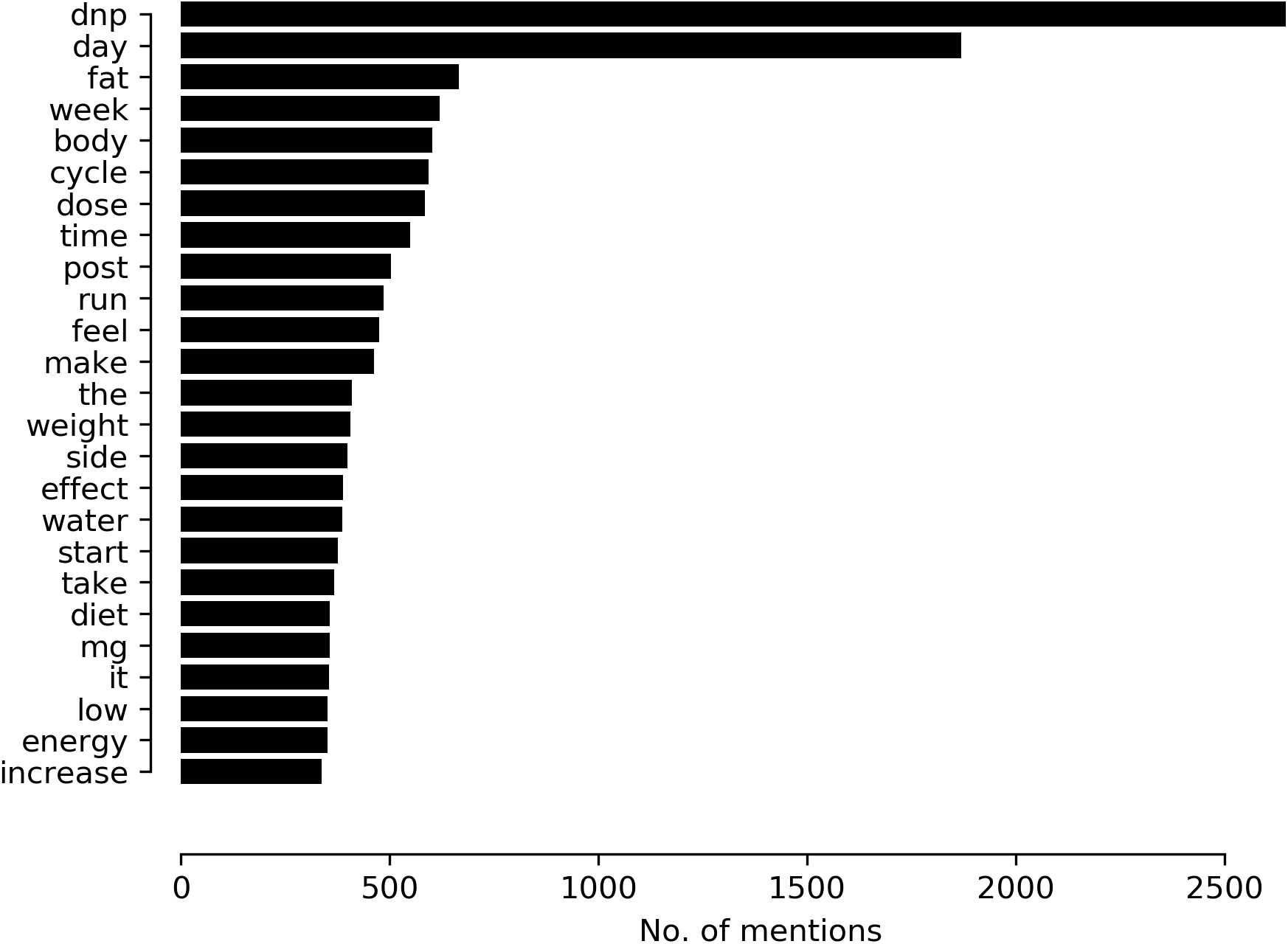
Twenty-five most frequently mentioned tokens. Y-axis shows token. X-axis counts number of mentions across all posts mentioning DNP.

### Relationship Between Dose and Effect

To better understand the dose-effect relationship of DNP we cross-tabulated words describing effects and doses in the subset of posts (n= 521) that mentioned DNP, dose, and effect (Figure 6). Most comments were made on doses between 100 mg - 300 mg, corresponding to 1-2 DNP pills. The most common dose at which comments explicitly stated that they experienced no side effects was 150 mg.

Two comments mentioned weight-dosing. One comment suggested 2 mg/kg was safe, but did not specify the duration of use or side effects. Another comment reported 30 mg/kg would be lethal, but did not provide the source for that information nor specify the duration of use.

**Figure 6.**
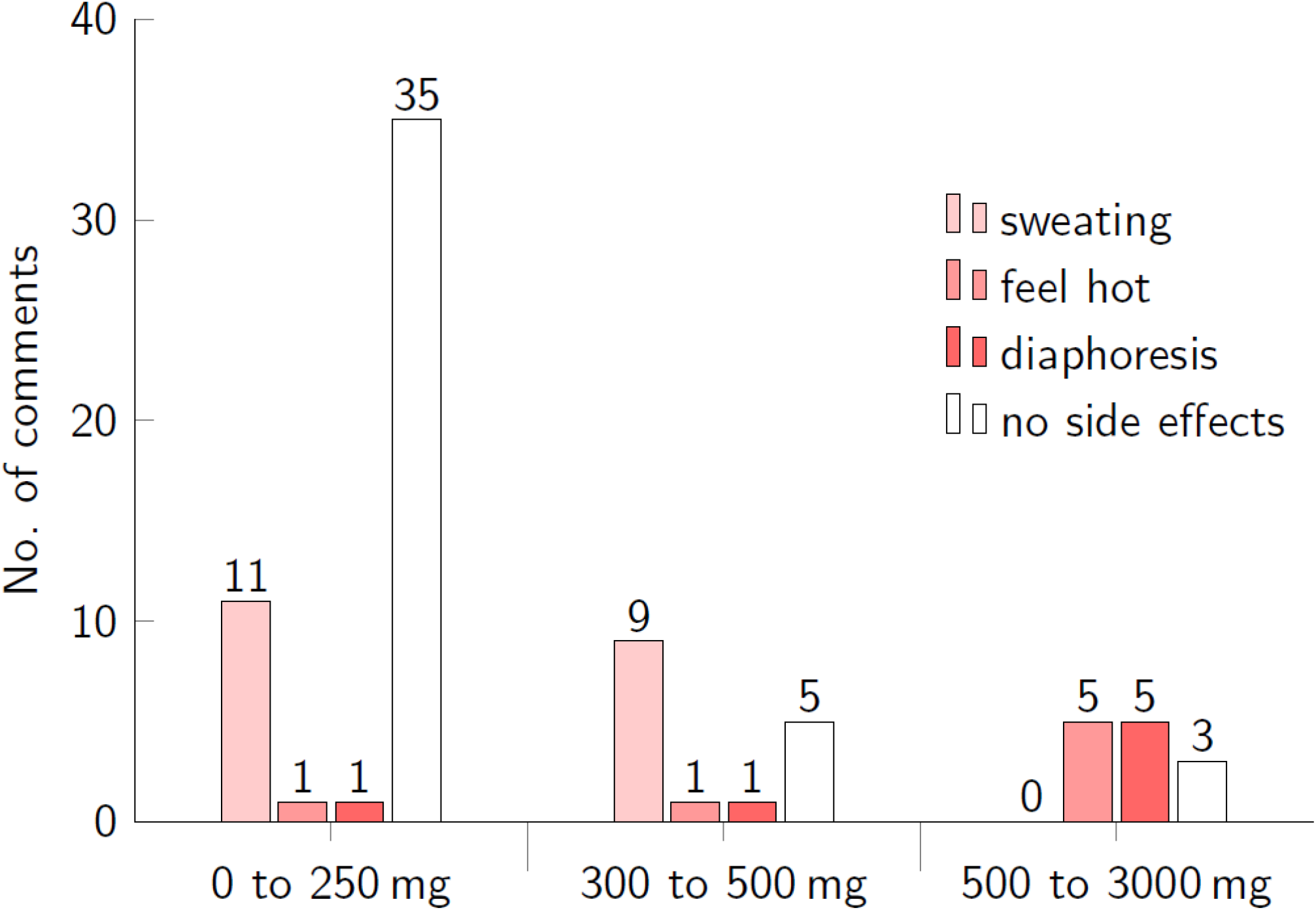
Reported effects of DNP. Each panel shows the number of mentions of an effect at different doses. X-axis is range of doses-in mg. Y-axis is number of mentions. Color of bar indicates symptom.

Figure 7 shows the distribution of duration of use mentioned in comments. The median duration was 10 ± 7 days (median ± interquartile range). The top three most commonly mentioned durations were 1, 4, and 20 days, corresponding to a single day trial as well as week and month trials with “on” and “off” periods. “On” and “off” periods referred to breaks in usage to avoid toxicity, similar to “sedation holidays” in intensive care units or medication holidays to limit adverse effects of selective serotonin reuptake inhibitors (SSRIs).

**Figure 7.**
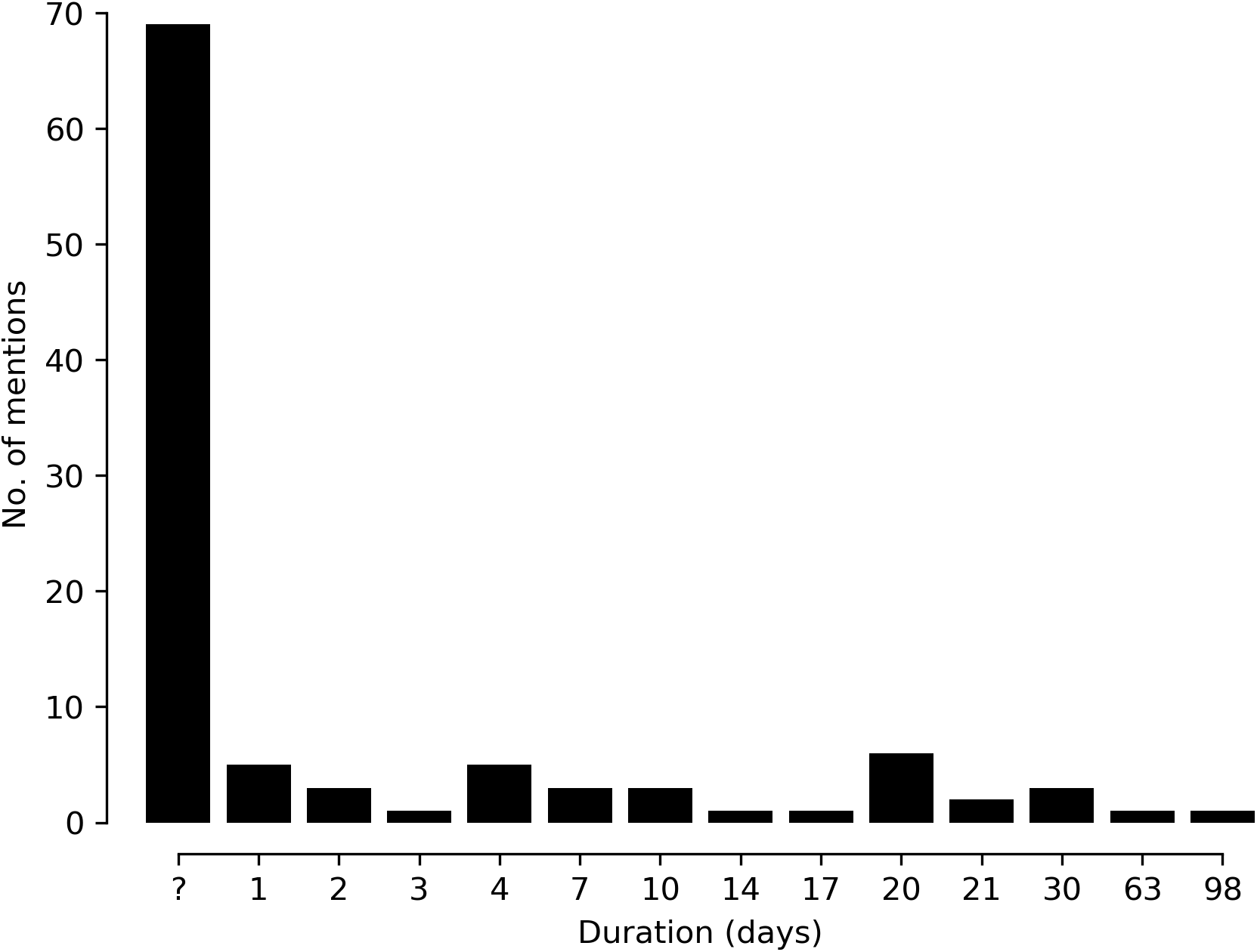
Distribution of duration of DNP cycle. X-axis shows number of comments mentioning the duration. Y-axis shows duration in days. The symbol “?” on the x-axis denotes a comment that mentioned DNP use but did not specify duration of DNP use.

### Analysis of Mentioned Coingestants

Anecdotal analysis of online bodybuilding forums suggest that DNP is consumed in between periods of anabolic steroid use. Co-ingestants mentioned online may be used to potentiate the intended effects of DNP, mitigate the unintended effects of DNP, or mitigate toxicity from anabolic steroid use. To identify the most significantly co-occurring coingestants, we calculated the Pearson correlation coefficient between the patterns of mention of DNP and all other substances. We used the Benjamini-Hochberg correction to account for testing multiple hypotheses on the same data set. Figure 8 shows the top 25 substances whose patterns of mention were significantly correlation with DNP’s pattern of mention. Fexofenadine (Claritin), and cetirizine (Zyrtec) were used to treat pruritus, although the posts seemed to confuse pruritus and paresthesias. Posts mentioning T3 (triiodothyronine) and DNP discussed the potential of DNP use to suppress endogenous T3 release and explained the effects of DNP use in terms of T3 use, including comparing DNP toxicity to thyrotoxicosis A synthetic version of T3 (liothyronine, brand name Cytomel) is also used to treat hypothyroidism.

**Figure 8.**
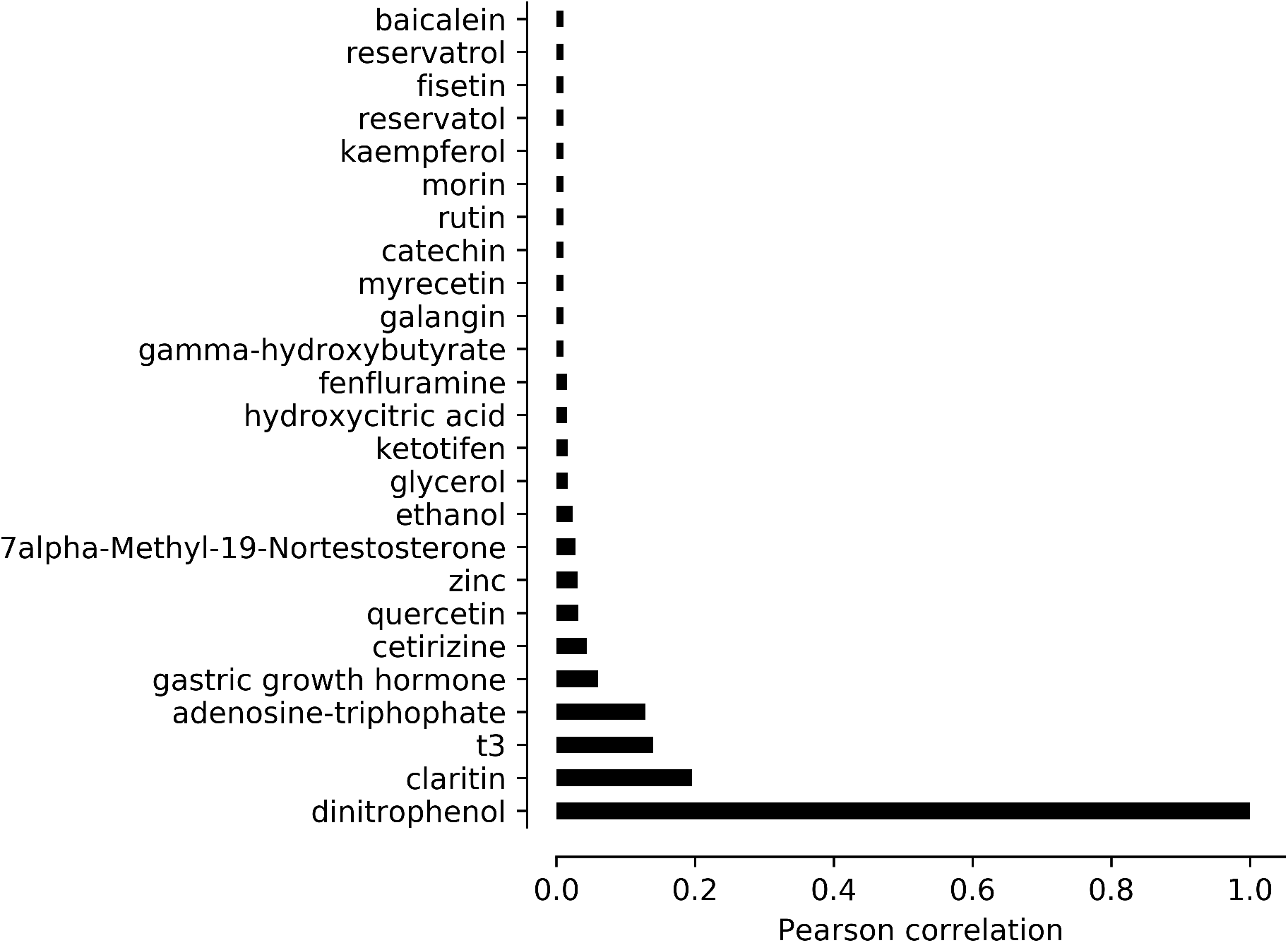
Substances with patterns of mention positively correlated with patterns of mention of DNP. Y-axis shows substance. X-axis shows Pearson correlation. Threshold for statistical significance r = 0.008.

Quercetin, zinc, and compounds myrecetin through baicalein (following the y-axis of Figure 8), are herbal supplements claimed to support the liver. Use of these herbal supplements was intended to mitigate the hepatotoxic effects of anabolic steroids. Mentions of tamoxifen and vitamin E, however, were also significantly negatively correlated with comments mentioning DNP. Tamoxifen is a SERM (selective estrogen receptor modulator) that could be used to mitigate the estrogenic effects of some steroids.

**Figure 9.**
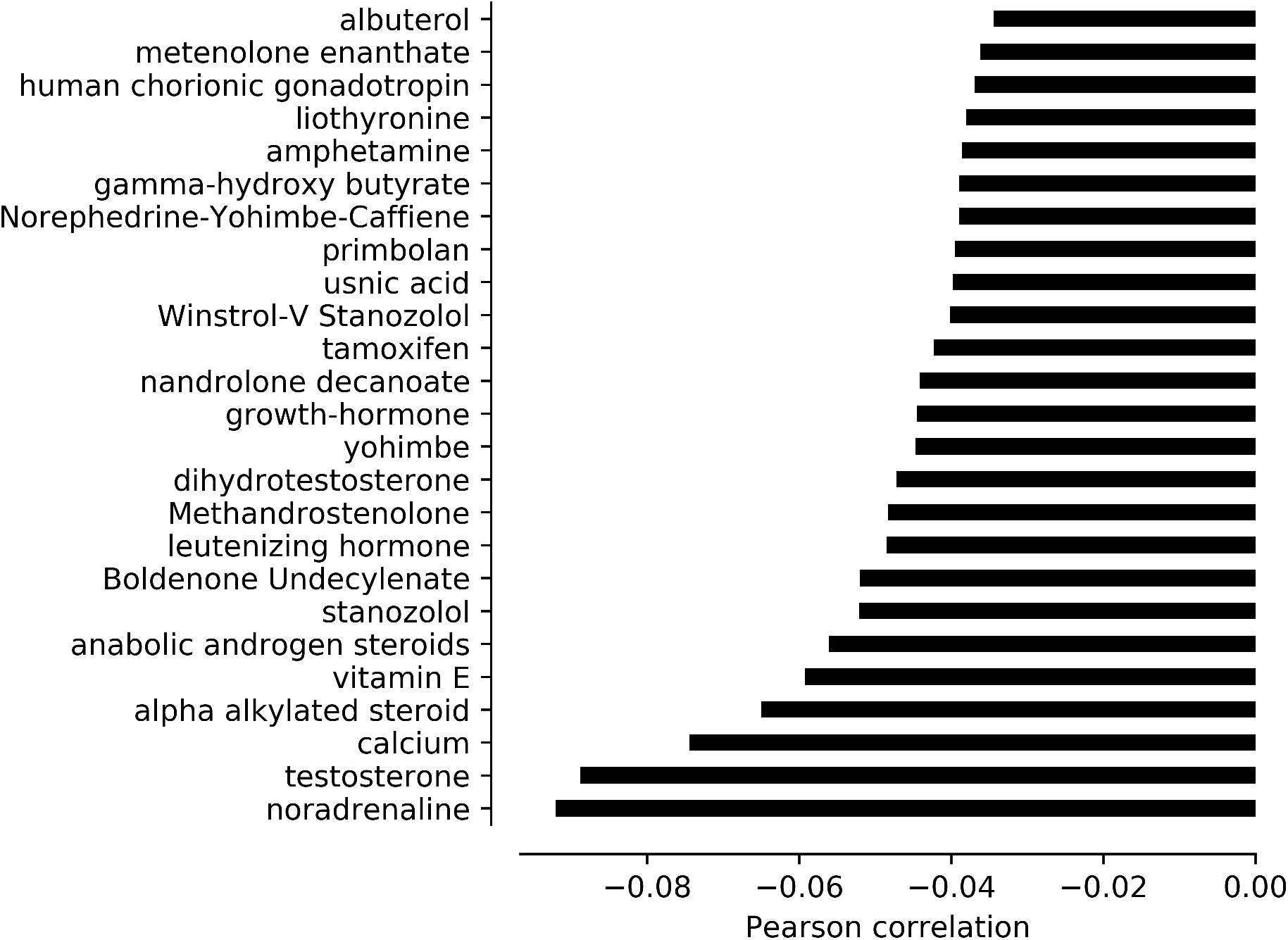
Substances with patterns of mention negatively correlated with patterns of mention of DNP. Y-axis shows substance. X-axis shows Pearson correlation. Threshold for statistical significance r = −0.001.

Figure 9 shows the substances whose pattern of mention was negatively correlated with the mention of DNP. The substances include stimulants (albuterol, liothyronine, amphetamine, caffeine, yohimbe, and noradrenaline). Supporting the anecdotal observation that DNP use occurred in between anabolic steroid use, the pattern of mention of DNP was significantly negatively correlated with the pattern of mention of any anabolic steroid (Figure 9).

## CONCLUSIONS

This is the first study to characterize the usage of DNP at nonlethal doses using social media. It estimated doses and effects consistent with prior literature, including reports with analytic confirmation of ingestion^7^. This study also revealed pruritus as a reported side effect at doses of 150-300 mg each day. Our study adds to the literature by supplying a thematic analysis of online discussions, identifying duration of use and co-ingestants, and identifying pruritus as a new unintended effect of sublethal DNP ingestion. Limitations to this study include lack of independent confirmation and reporting bias. Only 3/661 posts discussed sending a sample to a laboratory for GC-MS (gas chromatography-mass spectrometry) to verify composition. We presume that the rest used effect as a proxy for purity. The methods of production of DNP for human consumption are not regulated.

A unique limitation of online sources is the anonymity of the user. We suspect, based on the studies cited in the introduction, that the majority or users are adolescent and young adult males engaged in resistance training. Anonymity may facilitate discussion on illegal or sensitive topics, such as the consumption of a potentially effective but toxic substance banned for human consumption. We only considered unique accounts and did not analyze duplicate posts.

At room temperature DNP is a yellow powder, sold as a pill or powder. Users described measuring the powder out. The accuracy of commercial scales can vary from the order of 1 mg to 1 g, depending on the make. This range could contribute to inadvertent misdosing.

The presence of herbal and alternative supplements to “support the liver [and] metabolism” suggests an awareness of the adverse effects DNP can cause. Except for death and yellow discoloration, the forums did not mention longer-term sequelae previously noted in the literature such as cataracts, suggesting a bias to reporting short term effects. We found the median reported duration of use to be 10 days. 2,4-dinitrophenol (DNP) is an alluringly potent weight loss agent. Our work here suggests that doses of 150-300 mg, touted as “safe”, may still be associated with toxicity. This investigation also demonstrates the power of the internet to provide illuminating, if preliminary, results on topics with public health relevance not accessible through traditional means.

## Data Availability

All data are publicly available. All code for analysis is available on request.

